# Lateralized Circuitry for Verbal Fluency Changes After Subthalamic Nucleus Deep Brain Stimulation

**DOI:** 10.1101/2025.01.03.25319977

**Authors:** Min Jae Kim, Isabella Zuccaroli, Victor A. Del Bene, Sarah A. Brinkerhoff, Roy C. Martin, Joseph W. Olson, Matthew J. Nelson, Christopher L. Gonzalez, Vidyulata Kamath, J. Nicole Bentley, Robert T. Knight, Harrison C. Walker, Kelly A. Mills

## Abstract

**Background:** Subthalamic nucleus deep brain stimulation (STN-DBS) improves motor symptoms but can be associated with subtle cognitive changes, including declines in verbal fluency (VF). Network-based mechanisms underlying stimulation-induced fluency declines are not fully understood.

**Objective:** This study investigates how STN-DBS stimulation locations interact with brain connectivity patterns, and whether this differentially drives VF changes.

**Methods:** 20 patients with Parkinson’s disease and unilateral STN-DBS were analyzed. Electrodes were localized with estimates for volume of tissue activation (VTA). VTAs were seeded in normative functional and structural connectomes to identify connectivity profiles correlating with VF changes (R-maps). Additionally, amongst the fibers passing through VTAs, a two-sample *t*-test assigned fibers associated with VF improvement or decline.

**Results:** VF declines occurred following unilateral left but not right STN-DBS. R-maps revealed that improvements correlated with greater connectivity to prefrontal structures in right hemisphere such as inferior and superior frontal gyrus, whereas declines localized to structures on the left. Fibers filtered from right VTAs were associated with VF improvements. Fibers from left VTAs were associated with declines, but the gradient of fiber t-scores were distributed across an anterior-posterior axis ranging from supplementary motor area to paracentral lobule.

**Conclusion:** Unilateral STN-DBS interacts with distributed cognitive networks that display strong hemispheric lateralization. Beyond the effects of implant hemisphere, our findings suggest that connectivity between local STN subregions to larger whole-brain networks impact VF performance, in a manner that could yield more tailored approaches to electrode targeting and clinical programming.

## Introduction

Subthalamic nucleus deep brain stimulation (STN-DBS) is a widely accepted advanced therapy to control refractory motor symptoms from Parkinson’s Disease (PD). While motor improvements are typically substantial, measurable changes in neuropsychological function are reported in some patients. Studies report cognitive changes following STN-DBS, including declines in phonemic fluency, semantic fluency, executive function, and processing speed.^1^ For two types of verbal fluency (VF) - phonemic and semantic fluency – declines impact 20-40% of PD patients seen from early postoperative period.^2-4^

Verbal fluency declines following STN-DBS are likely associated with effects of both surgical implant^5^ and neurostimulation itself. In a trial incorporating sham stimulation, subjects were randomized to early or late device activation, and lead implant was associated with declines in phonemic and semantic VF. Phonemic fluency, however, worsened further upon device activation.^6^ Others suggest semantic verbal fluency declines with more anterior cortical entry points in the left hemisphere and upon passing through the caudate nucleus.^7, 8^ Previous work focused on active contact location and reports discrepant findings. For example, patients with more ventral contact activation in the STN experienced VF declines in some studies,^9, 10^ whereas more dorsal subregions in left STN led larger declines in another.^11^ A recent study further suggested that VF declines with left, but improves with right STN-DBS, highlighting a potential lateralization effect of stimulation.^12^

Beyond the local stimulation effect on the STN and nearby structures, how DBS engages brain networks connecting cortical and subcortical regions involved in language and speech production is largely unknown. Because VF requires distinctive cognitive components such as word selection and production that require recruitment of fronto-basal ganglia connections, minimizing stimulation effects on VF requires understanding the stimulation effect of STN-DBS at a network level. Understanding the “connectomic” effect of DBS on VF would help better guide accurate lead placement during surgery or steering stimulation current during postoperative follow-up periods.

Here we used data from a unique cohort of patients who underwent unilateral STN-DBS^8^ to investigate how stimulation locations interact with brain connectivity patterns, and whether this differentially drives VF changes. We hypothesized that verbal fluency declines are associated with stimulation sites with connectivity to cortical regions with known roles in verbal processing and language production.

## Materials and Methods

### 1. Patient Recruitment and Clinical Evaluation

Twenty people with PD underwent unilateral STN-DBS directional electrode implantation (Vercise Directed, Boston Scientific) as part of a prior study.^8^ We analyzed data from participants who had the requisite neuroimaging and clinical outcomes and reviewed longitudinal letter fluency and MDS-Unified Parkinson’s Disease Rating Scale (MDS-UPDRS) Part III scores. Letter VF (FAS or CFL, alternating between baseline and post-DBS) scores were normalized to T-scores, adjusting for demographic factors (age, race, sex, education).

Pairwise preoperative and 2-month postoperative VF and MDS-UPDRS scores were compared using Wilcoxon signed-rank test with statistical significance set at p < 0.05. All graphical and statistical analyses, including neuroimaging processing, were performed using MATLAB 2022b (MathWorks, MA) and Prism 9 (GraphPad Software, CA).

### 2. DBS Electrode Localization

Unilateral DBS electrodes were localized by co-registering intraoperative CT sequences to pre-operative T1/T2 MRI sequences in the open-source Lead-DBS toolbox^13^ and then normalized to 2009 ICBM MNI space using Advanced Normalization Tools. Electrodes were reconstructed based on postoperative CT artifacts using a refined TRAC/CORE method,^14^ and the directionality of contacts was predicted via the DiODe algorithm both implemented within the Lead-DBS platform.^15^

Based on the active contact and respective stimulation amplitude, an electric field model centered around the reconstructed electrode contact was produced in the SimBio/Fieldtrip pipeline based on neighboring tissue conductivities.^14^ Finally, as per previous investigations,^16-18^ a binary volume of tissue activation (VTA) was produced by thresholding the electrode field model to contain only regions where the field strength was higher than 0.2 V/mm.

### 3. Mean Effect Image (MEI) Generation

We generated mean effect images (MEIs) to understand how different stimulation volumes targeting the STN relate to changes in verbal fluency and motor function. DBS-induced percent change in MDS-UPDRS and VF T-scores were assigned to voxels corresponding to left and right hemisphere VTAs. These weighted VTAs were then averaged across participants yielding MEIs specific to changes in both motor and verbal fluency function.^19, 20^ Regions with positive MEI values suggest a cohort-level increase in verbal fluency or motor function, whereas those with negative values reflect fluency declines. We did not specify a statistical threshold for probabilistic stimulation maps because of the limited sample size, particularly given further division into subgroups based on implant hemisphere.

### 4. Structural Connectivity and Fiber-filtering Analysis

To characterize the association between VF changes and structural connections shared between VTAs and other regions, we adopted a normative structural connectome (PPMI-85) with 1,700,000 fibers across 85 PD patients generated through the Parkinson’s Progressive Markers Initiative (PPMI).^21^ By seeding the connectome with individual VTAs, a structural connectivity (SC) profile denotes the number of fibers passing through VTAs to whole-brain voxels. To identify fibers strongly associated with VF changes, SC profiles of each patient were spatially correlated with respective VF changes, resulting in a voxel-wise “R-map”. Voxels with positive values in the R-maps represent specific regions where more fibers passing through the VTAs to those regions are associated with VF improvement, and negative values with decline.

To test the hypothesis that our *a prio*ri regions of interests are implicated in VF, mean SC across 166 cortical and subcortical structures were derived from the Automated Anatomical Labelling Atlas 3 (AAL3)^22^ were correlated with VF changes. Our *a priori* regions were inferior frontal gyrus subregions (IFG), lateral orbital frontal cortex (OFC), supplementary motor area (SMA), and mesial temporal structures such as amygdala and hippocampus that have been shown to play significant roles in verbal processing and speech production. Statistically significant regions associated with VF changes were identified post-hoc after correcting for the false discovery rate.

Finally, fiber-filtering of individual fibers from the connectome was adopted to test whether targeting a certain fiber is more likely to result in VF decline or improvement as well as the magnitude of VF change. Following the methods from similar network mapping studies,^23-25^ fiber-filtering was conducted such that for each fiber, VTAs of the patient cohort were classified into those connected to the fiber and those not connected. Across these two groups, their respective VF changes were compared using a two-sample t-test, resulting in a t-value assigned to each fiber.

The magnitude and sign of the fiber t-value corresponds to how strongly the stimulation of that fiber is associated with VF changes as well as the relative direction of changes (VF decline or improvement), respectively. To reduce potential type II and type I statistical errors, only fibers connected to more than 10% but less than 90% of total VTAs for each side were considered for analysis. For each fiber, after performing a two-sample t-test between VF changes in unconnected and connected VTAs, only fibers assigned with the top 20% and the bottom 20% of t-scores were considered for subsequent analysis on each side of the stimulation.

### 5. Functional Connectivity Analysis

While the effect of STN-DBS can partially be explained by monosynaptic, antidromic connections from STN to cortical regions via the hyperdirect pathway,^26^ the mechanism of action of DBS’s effect may also include alteration in signaling through the indirect cortico-basal ganglia-thalamo-cortical pathway, a polysynaptic pathway better represented by functional connectivity.^27^ To evaluate how functional connectivity (FC) profiles are associated with VF changes, we produced FC maps specific to each VTA by seeding VTAs into a normative functional connectome (GSP-1000) produced from fMRI scans across 1000 healthy adults through the Brain Genetics Superstruct Project.^28^ FC maps then underwent Fisher Z-transformation to normalize fMRI BOLD correlation signals across whole-brain voxels. Similar to R-map analysis for SC profiles, each normalized FC profile was spatially correlated with VF changes to produce a voxel-wise functional R-map.

### Data Sharing

The data that support the findings of this study are available upon reasonable request to the corresponding author, KAM. The data are not publicly available due to information that could compromise the privacy of patient subjects.

## Results

### 1. Demographics and Clinical Information

Eleven of the 20 patients (55%) had left unilateral STN-DBS and 15 (75%) were right-handed (**Supplementary Material 1**). Based on the electrode reconstructions visualized in **Figure 1 (top row)**, unilateral electrodes on either side were localized within the posterolateral sensorimotor region of the STN.

**Figure 1.**
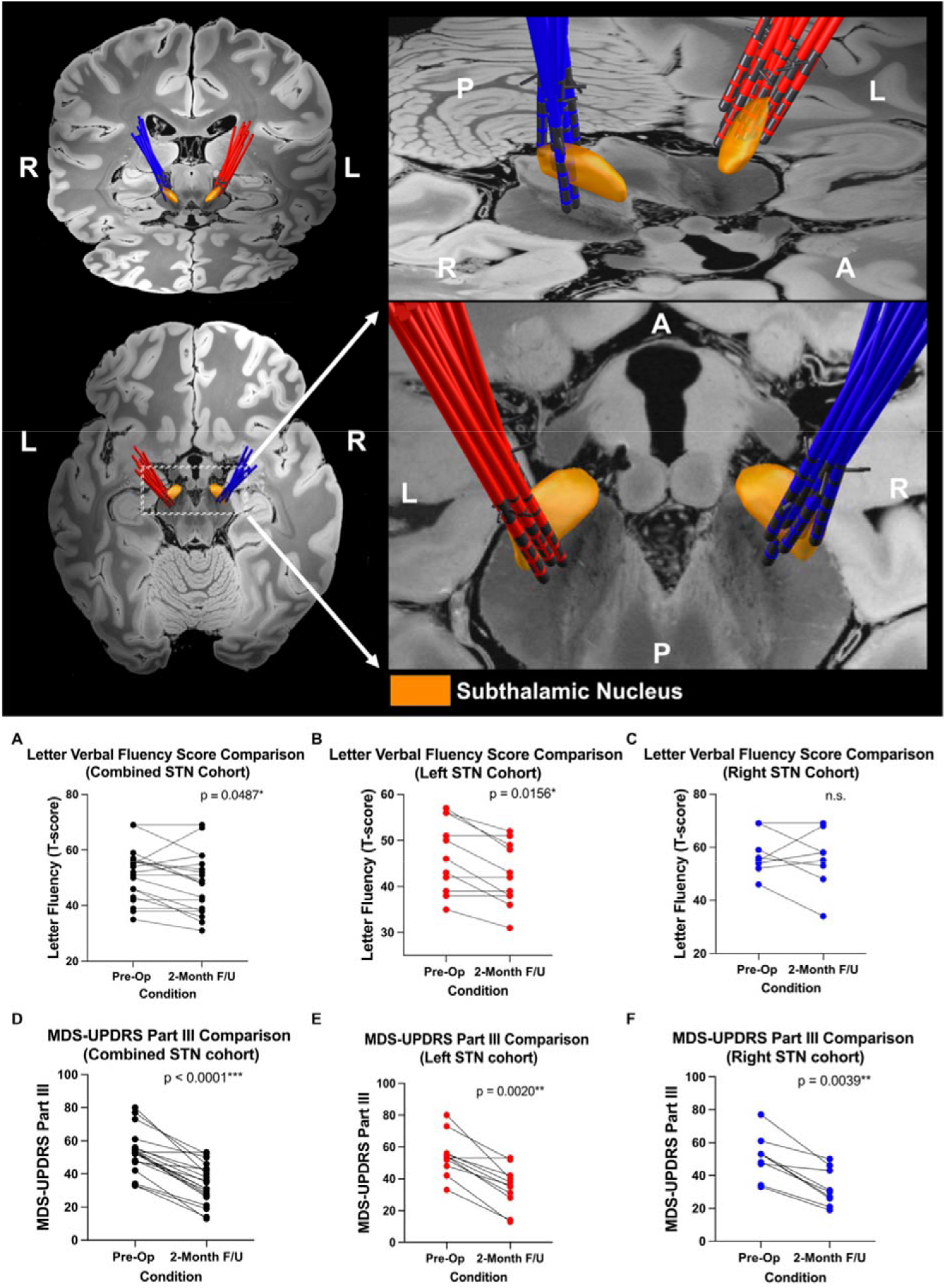
DBS Electrode Visualization and Postoperative Clinical Changes. Unilateral STN-DBS electrodes (right = blue, left = red) were localized and shown relative to STN (orange) from DISTAL basal ganglia atlas (top row). A = Anterior, P = Posterior, L = Left, R = Right. Postoperative Changes in VF and MDS-UPDRS Part III Scores (bottom row).

### 2. PD Motor and Verbal Fluency Changes

Patients who underwent left STN-DBS displayed significantly lower pre-operative verbal fluency versus those who underwent right hemisphere implants (left=46.6±3.7, right=57.1±2.5,p=0.0209) as shown in **Figure 1A-C**. No significant VF changes were observed in right STN-DBS patients at a group level, but 4 patients had higher VF scores relative to preoperative performance and 4 patients had lower postoperative VF scores relative to preoperative baseline. The left STN-DBS cohort showed a -8.6±2.2% change, and the right STN-DBS cohort showed a -2.4±5.2%, suggesting that left-side stimulation is associated with more prominent VF decline even when accounting for baseline VF performance.

MDS-UPDRS Part III motor scores were significantly improved for both right (-37.3±5.7%, p=0.0039) and left STN-DBS (-36.3±4.7%,p=0.002) cohorts relative to preoperative conditions **(Figure 1D-F)**.

### 3. Mean Effect Image (MEI) of Verbal Fluency and PD Motor Changes

From the left STN-DBS cohort, stimulation sites associated with most VF decline were localized within the posterior STN in its dorsal-medial border (**Figure 2A Top)**. For right STN-DBS cohort, stimulation sites associated with most VF improvement were localized within the posterior STN in its ventral-lateral border (**Figure 2A Bottom**). For both right and left STN-DBS cohorts, greater improvement in MDS-UPDRS was associated with stimulation of the posterior-lateral region of the STN.

**Figure 2.**
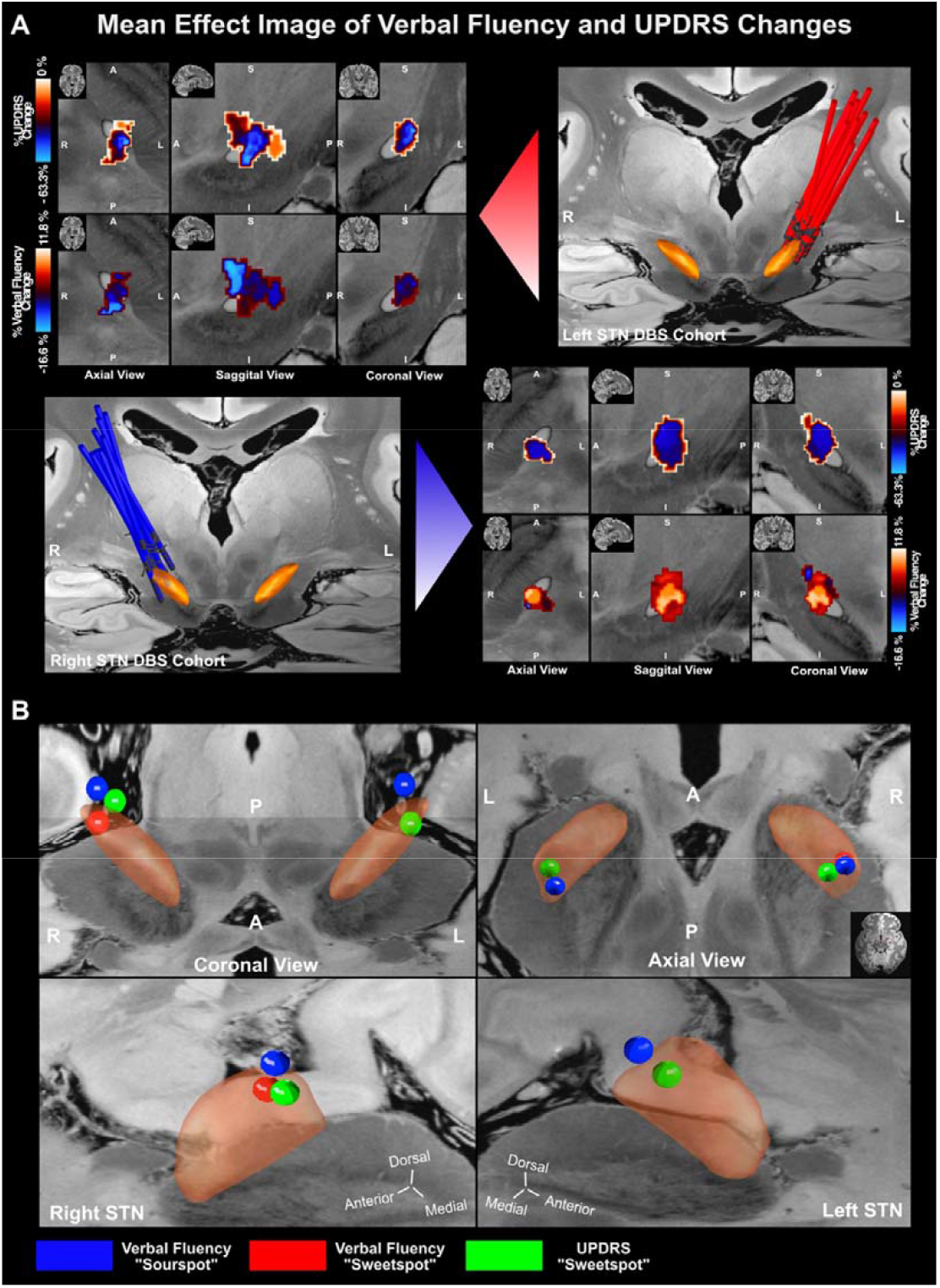
STN Mean Effect Image (MEI) of VF and MDS-UPDRS Part III Scores. (A) Spatial gradient of VF and MDS-UPDRS Part III changes across the STN for left (top row) and right (bottom row) STN-DBS. View of MEI is shown at x = -14.1mm, y = -14.7mm, z = -5.1mm. (B) Centroid Coordinates of VF and MDS-UPDRS Changes. VF “sweetspot” was defined by centroid coordinates of top 20% of voxels, and VF “sourspot” was defined by centroid coordinates of bottom 20% of voxels comprising VF MEI for each side of stimulation. Because the maximum voxel value of the left VF MEI was 0, VF “sweet spot” was not produced as all voxels represented a global decline of VF. Centroid coordinates of bottom 20% of voxels of MDS-UPDRS MEI is shown in green. R = Right, L = Left, A = Anterior, P = Posterior, R = Right, S = Superior, Inferior.

To more precisely localize stimulation sites associated with VF change, MEIs were restricted to 20% of voxels that showed the strongest correlation with clinical outcomes. **(Figure 2B)**. The centroid coordinates of the top 20% of VF MEI voxels and bottom 20% of MDS-UPDRS MEI voxels are defined as the “sweet spot” and the centroid coordinate for bottom 20% of VF MEI voxels as “sour spot”.

For the right side, the VF “sweet spot” was localized at x= 4.21mm, y=-13.95mm, z=-5.97mm (average VF change = 8.67±1.46%) and VF “sour spot” at x=14.22mm, y=-14.45mm, z=-3.17mm (average VF change = -9.61±1.40%). MDS-UPDRS “sweet spot” was at x=12.60mm, y=-15.20mm, z=-4.65mm (average MDS-UPDRS change=-46.75±0.67%). For the left hemisphere, the VF “sour spot” was at x=-13.55mm, y=-17.21mm, z=-4.20mm (average VF change=-14.63±0.37%) and MDS-UPDRS “sweet spot” was at x=12.60mm, y = -15.20mm, z=-4.6483mm (average MDS-UPDRS change=-46.75±0.67%).

### 4. Structural Connectivity Profiles Associated with VF Changes

Based on R-map shown in **Figure 3 (upper row)**, increased connectivity between VTAs with right-side cortical structures including amygdala (r=0.548,p=0.012), lateral OFC (r=0.602,p=0.0049), medial OFC gyrus (r=0.467,p=0.038), temporal pole of middle temporal gyrus (r=0.55,p=0.013) were significantly correlated with VF improvement. Structural connectivity with other hypothesized regions of interest was not significantly correlated with VF improvement, but there was a trend for a correlation between VF improvement and connectivity with the pars orbitalis (r=0.41,p=0.07).

**Figure 3.**
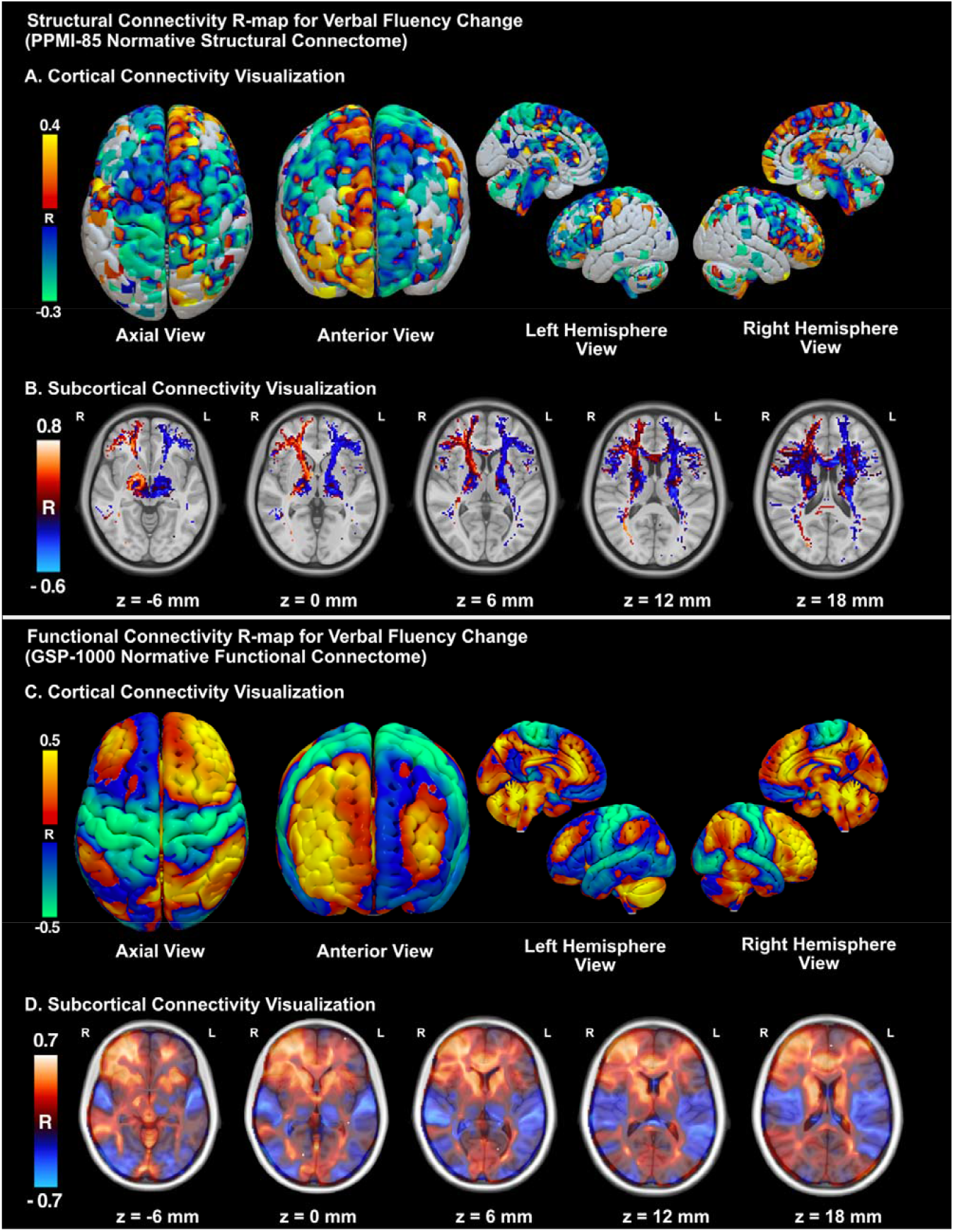
Structural and Functional R-map of VF Changes. For structural connectivity (upper row), spatial correlation “R” values between number of fibers passing through VTA and VF changes are shown on A) cortical surface and (B) subcortical axial view. For functional connectivity (lower row), spatial correlation “R” values between functional connectivity relative to VTA and VF changes are shown on (C) cortical surface and (D) subcortical axial view. R = Right. L = Left. Higher warm color intensity (yellow-red) = higher connectivity is correlated with VF improvement. Higher cool color intensity (blue-green) = higher connectivity is correlated with VF decline.

For the left hemisphere, there was not a significant correlation between VF decline and structural connectivity with the hypothesized structures: left IFG pars opercularis (r=-0.011,p=0.96), pars orbitalis (r=-0.23,p=0.33), pars triangularis (r=-0.25,p=0.28), lateral OFC (r=-0.067,p=0.78), SMA (r=-0.23,p=0.33), and amygdala (r=-0.087,p=0.71).

### 5. Functional Connectivity Profiles Associated with VF Changes

As shown in **Figure 3 (lower row)**, increase in functional connectivity to right-sided regions including, but not limited to, anterior cingulate cortex (r=0.45,p=0.045), IFG pars orbitalis (r=0.45,p=0.048), IFG pars triangularis (r=0.47,p=0.037), middle frontal gyrus (r=0.48,p=0.03), and super frontal gyrus (r=0.48,p=0.034) was significantly correlated with postoperative VF improvement.

On the other hand, an increase in functional connectivity with left side structures as left amygdala (r=-0.52,p=0.019) and paracentral lobule (r=-0.47,p=0.036) extending to primary motor and somatosensory cortex were significantly correlated with postoperative VF decline. Correlations between VF decline and functional connectivity with our hypothesized regions of interest were not observed: IFG pars opercular (r=-0.16,p=0.49), pars orbitalis (r=-0.074,p=0.75), pars triangularis (r=-0.07,p=0.74), lateral OFC (r=-0.12,p=0.62), SMA (r=-0.22,p=0.34), and hippocampus (r=-0.26,p=0.26).

### 6. Lateralization of White Matter Fibers Associated with VF Changes

For the right side, a total of 1035 tracts passed through the right STN VTAs, and their mean fiber T-score was 2.148. 97.6% (1010) of tracts were positive (t-score=2.261 ±0.003), and 2.4% (25) were negative (t-score=-2.410±0.000). Strongly positive fibers associated with VF improvement tended to run in the anterior-lateral border, while strongly negative fibers associated with VF decline through the posterior-medial border of the posterior STN **(Figure 4A-B)**.

**Figure 4.**
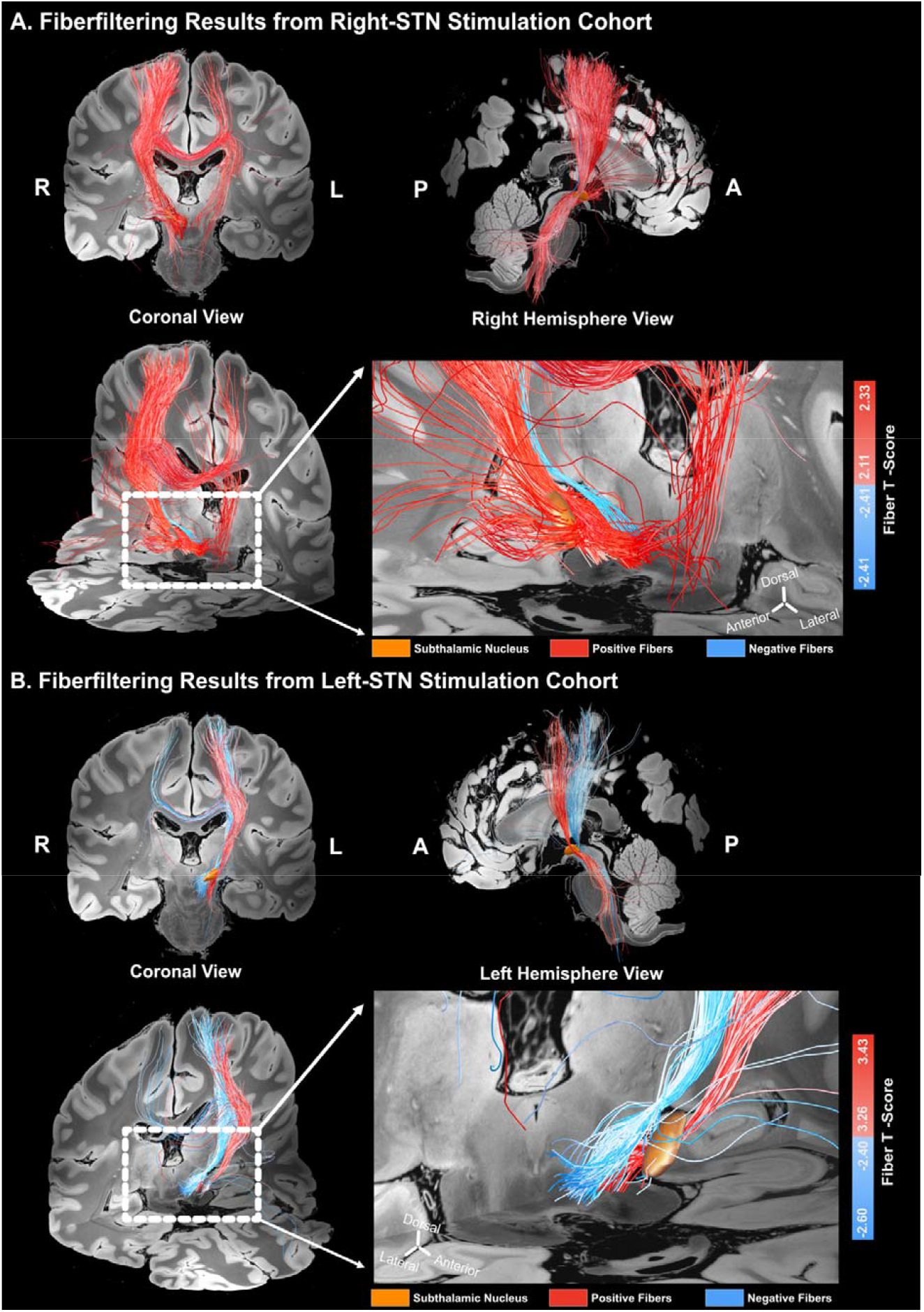
Fiber tracts Associated with VF Changes of Unilateral STN-DBS Cohort. For each right (A) and left (B) side stimulation cohort, filtered fibers are visualized in coronal, sagittal, and oblique views. Red fibers are denoting fibers assigned with positive (VF improvement) t-score, and blue with negative (VF worsening) t-score. Only fibers with only top and bottom 20% t-scores were visualized. R = Right, L = Left, A = Anterior, P = Posterior.

On the other hand, total 299 tracts transverse through the left STN VTAs with mean t-score of -0.5534 as shown in **Figure 4C-D**. Because all patients either had a decline of VF or no change for left side cohort (i.e no improvement), strongly positive fibers represent fibers associated with relatively less VF decline, and strongly negative fibers with relatively more VF decline. From 299 fibers, 33.1% of fibers were positive fibers (mean t-score=3.349±0.008), and 66.9% negative fibers (mean t-score=-2.485±0.007). Similar to trend shown in right STN VTAs, positive fibers corresponding to less VF decline passed through more anterior-lateral border, and negative fibers corresponding to more VF decline through posterior-medial border of the posterior STN.

In a combined cohort, a distinctive connectivity pattern across hemispheres associated with VF changes was found as shown in **Figure 5**. Fibers from right VTAs were predominantly labeled with positive t-scores for VF improvement. For left side VTAs, two clusters of fiber bundles were produced, where more anterior-division fibers and posteriorly-division fibers were associated with VF improvement and VF decline, respectively. Furthermore, for both right and left side VTAs, cortical projections of positive fibers tend to localize more anteriorly to supplementary motor area while negative fibers to more posterior regions in primary motor and somatosensory cortex **(Figure 5B)**.

**Figure 5.**
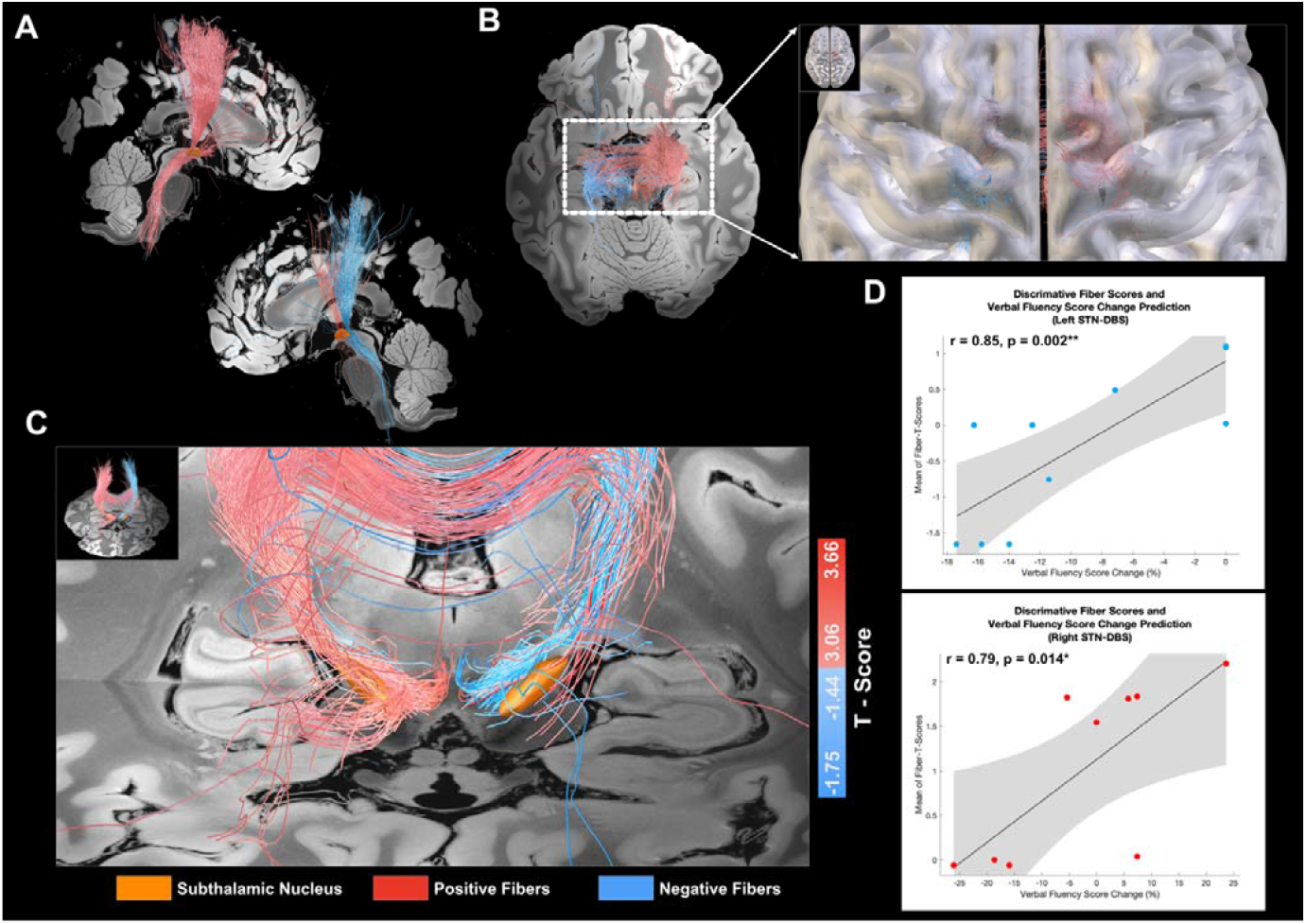
Fiber tracts Associated with VF Changes of Combined STN-DBS Cohort. Filtered fibers are visualized in (A) sagittal and (C) coronal view. Red fibers denote fibers assigned with positive t-score, and blue with negative t-score. Only fibers with only top and bottom 20% t-scores were visualized. (B) Fiber track connection to the paracentral lobule and dorsolateral prefrontal cortex. (D) Correlation between VF score change and mean fiber t-scores for each patient from left (top) and right (bottom) STN-DBS cohort.

Finally, mean of t-scores for all the fibers passing through each VTAs were correlated with respective VF changes separately for left and right VTAs. For both left (r=0.85,p=0.002) and right (r=0.79,p=0.014) sided VTAs, fiber t-scores were strongly associated with VF changes (**Figure 5D)**.

## Discussion

In our study, we investigated the location of stimulation sites and circuitries associated with VF change following unilateral STN-DBS of either hemisphere in a cohort of patients that, on average, experienced a decline in VF in a 2-month follow-up period. Having established that the presence of VF decline in our combined group and the left hemisphere group is consistent with findings on VF in bilateral STN-DBS cohorts from other studies, and that the more mixed outcomes in our right hemisphere aligned with prior studies^29^, we found that VF change may be differentially modulated by the location of stimulation sites relative to the STN and surrounding structures. Furthermore, there may be a differential cortical connectivity pattern across the hemispheres associated with VF decline or improvement. Structural and functional connectivity profiles seeded by stimulation volumes further informed how intervening in a lateralized circuitry involving IFG, dorsolateral prefrontal cortices (PFC), and mesial temporal regions has an effect on VF changes.

The MEI or “sweet/sour spot” analysis of VF demonstrated a different spatial distribution of VF changes between right and left STN mainly across the dorsomedial and ventrolateral border of STN. Also supporting this gradient of effect on VF, our fiber-filtering results illustrate an anterior-posterior division of fiber bundles associated with the degree and directionality of VF changes for both hemispheres. For right-side STN stimulation, anterior bundle (positive fibers) extending towards SMA and prefrontal cortices was more associated with VF improvement, and the posterior bundle (negative fibers) headed toward somatosensory cortices with VF decline. For left-side stimulation, a more posterior bundle was associated with more VF decline. To better contextualize relative stimulation sites associated with VF and MDS-UPDRS changes, centroid analysis in **Figure 2** proposes respective “sour spot” and “sweet spot” in the STN. Because VF “sour spots” were located posterior-dorsomedial portion in bilateral STN, this may explain the findings that more VF decline was observed in the dorsal subregion of left STN in a previous report.^8, 30^. Furthermore, because motor sweet spots for PD are also recognized to be in the posterior dorsal portion of the STN,^12, 31, 32^ the significant overlap of MEI of VF and MDS-UPDRS has implications for the placement of directional contacts in this region to facilitate VTA shaping that maximizes motor benefits while minimizing dorsomedial spread to the MEI of VF “sour spot”.

While the spatial gradient of VF changes in the subthalamic area remains consistent across the two hemispheres, the magnitude of VF changes varies such that MEI of the left STN are predominantly composed of VF decline (-16.6% to 0%), and right STN with a combination of VF decline and improvement (-15.7% to 15.5%). If this hemispheric effect is validated in other cohorts, it could be considered during the consultation of patients regarding the risk of verbal fluency changes with DBS as it may differ regarding the hemisphere targeted. Also, this bilateral overlap of the MEI for VF decline and MDS-UPDRS improvement that, in the left hemisphere, is not offset by improvement seen with stimulation of more anterior fibers as in the right hemisphere may impact the decision to implant unilateral (right) or bilateral STN leads in cases who would be particularly sensitive to VF decline or where baseline VF function is already impacting quality of life, especially where left-sided motor symptoms predominate.

For functional (FC) and structural (SC) profiles associated with VF changes, there was hemispheric lateralization of networks associated with VF changes. Specifically, increased structural connections to right lateral OFC and middle temporal gyrus, as well as increased functional connections to right IFGs subregions and dorsolateral PFC, are associated with VF improvement. This connectivity pattern overlaps with a previously established network involved in VF, including a connection between the left inferior frontal gyrus (IFG) and superior frontal gyrus regions including SMA through the frontal aslant tract. The left IFG plays a role in both phonemic and semantic fluency, whereas the posterior bilateral IFG is more involved in phonemic retrieval.^33, 34^ The left IFG is connected to pre-SMA/SMA for motor selection and speech execution.^35, 36^ Past studies have identified a hyperdirect pathway between the STN and SMA, as well as STN and IFG, ^37^ which suggests the modulatory role of STN on VF through projections to the prefrontal and premotor cortices.

More ventromedial prefrontal structures such as in lateral OFC where clusters of Brodmann’s Area (BA) 47 are located are reported to be highly activated during both phonemic and semantic VF performances ^38^. Left rolandic operculum activity (IFG pars operacularis) is also shown to be involved in predominantly phonemic VF over semantic VF ^39^. Lastly, some evidence draws attention to the role of left medial temporal regions in phonemic VF ^39, 40^, but these regions are generally characterized to be involved more in semantic VF ^41-44^.

Our observation that right hemispheric stimulation has a substantial impact on VF performance may seem contradictory to our current understanding of VF function lateralization to the left hemisphere. However, we only analyzed patients who were implanted unilaterally in either the right or left hemisphere since our goal was to be able to parse the individual hemispheric effects on VF. As such, the results of this analysis do not speak to the combined effect of bilateral stimulation at the patient level. When all of our patients are analyzed as a combined group, those with left STN stimulation experienced a larger degree of VF decline than patients with right STN stimulation, suggesting that VF decline is a consequence of stimulating the ipsilateral, language-dominant hemisphere in a group who is presumed to be mostly left language dominant given the predominance of right-handedness.

One potential explanation might be a relatively higher preoperative VF performance of patients who underwent right STN-DBS than patients with left STN-DBS, but a prior analysis found that adjusting for preoperative VF performance did not change the differential effect of STN DBS on VF by hemisphere.^8^ However, the variance in VF performance within each hemisphere group differed and this could affect the analysis: standard deviation of VF changes on the right STN-DBS cohort was 15.65%, and the left STN-DBS cohort of 7.34%.

Consequently, higher variability of VF changes for the right STN-DBS may have augmented the correlation results with the structural and functional connectivity measures of each patient since there would be more parametric space to drive associations in the right hemisphere data set.

Several limitations exist in this study. First, this is a retrospective cross-sectional study investigating the effect of DBS on VF at 2-month postoperative period. Patients were not randomized to receive either right or left STN-DBS and thus inherent differences in VF based on which hemisphere is more affected (and thus, targeted with surgery) could underly some of the differences in this study. However, our findings of a gradient of effect on VF in the right hemisphere show that stimulation location and network engagement do contribute an independent effect beyond the hemisphere of stimulation. Also, because acute VF changes from immediate DBS-ON/DBS-OFF trials were not measured, our results may potentially be confounded by levodopa equivalent daily dose (LEDD). However, results from randomized control trials concluded that LEDD are not related to VF decline, and were not considered to perform any covariate analysis.^45, 46^

Second, we utilized a normative connectome to study relevant circuit nodes in VF that was derived from a large, population-based cohort rather than using patient-specific tractography results. Because structural and functional connectivity properties in PD patients may be altered compared to healthy controls,^47-49^ and may be variable within a PD patient cohort, such disease-specific features may limit the generalizability of our findings. Furthermore, previous study have shown an continual improvement of VF following right STN-DBS over time.^8^ With the dynamic nature of VF changes across longer postoperative timepoints, our analysis may not capture and reflect dynamic changes of underlying functional networks recruited. However, similar to other network-mapping studies in PD,^21, 50^ our goal of adopting normative connectome was to identify neural substrates commonly involved in VF function and its isolated changes from the stimulation effect. Nevertheless, future investigation is needed to study the VF response of DBS in the context of patient-specific fMRI or dMRI sequences.

## Conclusion

In this study of PD patients who underwent unilateral STN-DBS, the magnitude and direction of phonemic VF changes were not only dependent on stimulation sites in the anterolateral– posteromedial axis of STN, but also the hemisphere of stimulation. Our results suggest potential network-based mechanisms for the VF change commonly seen in PD patients undergoing STN-DBS and could be considered in counseling patients on the risk of VF changes with various surgical approaches to STN DBS or in targeting and stimulating subregions of the STN.

## Author Roles

1. Research project: A. Conception, B. Organization, C. Execution;
2. Statistical Analysis: A. Design, B. Execution, C. Review and Critique;
3. Manuscript Preparation: A. Writing of the first draft, B. Review and Critique;

M.J.K: 1A-C, 2A-B, 3A-B

I.Z: 2A-B

V.A.D: 2C,3B, 3B

S.A.B: 2B-C

R.C.M: 2B-C

J.W.O: 2B-C

M.J.N: 2B-C

C.L.G: 2B-C

V.K: 2B-C, 3B

J.N.B: 2B-C, 3B

R.T.K: 2B-C, 3B

H.C.W: 2C,3B

K.A.M: 1A-B,3B

---

**Supplementary Material 1:**
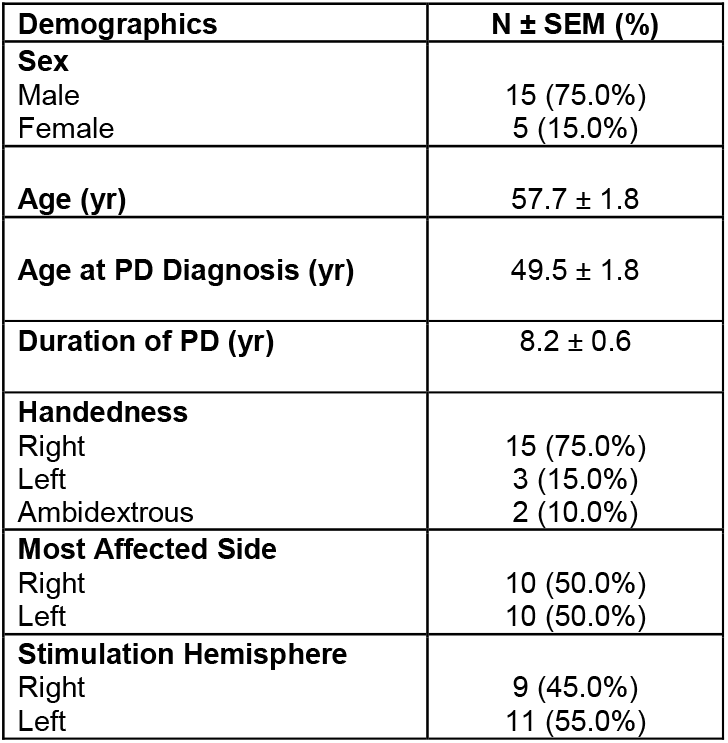
Patient Demographic and Clinical Information.

